# DiagnosisQA: A semi-automated pipeline for developing clinician validated diagnosis specific QA datasets

**DOI:** 10.1101/2021.11.10.21266184

**Authors:** Shreya Mishra, Raghav Awasthi, Frank Papay, Kamal Maheshawari, Jacek B Cywinski, Ashish Khanna, Piyush Mathur

## Abstract

Question answering (QA) is one of the oldest research areas of AI and Computational Linguistics. QA has seen significant progress with the development of state-of-the-art models and benchmark datasets over the last few years. However, pre-trained QA models perform poorly for clinical QA tasks, presumably due to the complexity of electronic healthcare data. With the digitization of healthcare data and the increasing volume of unstructured data, it is extremely important for healthcare providers to have a mechanism to query the data to find appropriate answers. Since diagnosis is central to any decision-making for the clinicians and patients, we have created a pipeline to develop diagnosis-specific QA datasets and curated a QA database for the Cerebrovascular Accident (CVA). CVA, also commonly known as Stroke, is an important and commonly occurring diagnosis amongst critically ill patients. Our method when compared to clinician validation achieved an accuracy of 0.90(with 90% CI [0.82,0.99]). Using our method, we hope to overcome the key challenges of building and validating a highly accurate QA dataset in a semiautomated manner which can help improve performance of QA models.

## 1 Background & Summary

Question Answering (QA) is a primary downstream task for natural language processing. QA systems for databases were a particularly popular research topic in the 1980s and early 1990s[1]. Currently, Question Answering (QA) is a benchmark Natural Language Processing (NLP) task where models predict the answer for a given question using related documents, images, knowledge bases. Biomedical QA(BQA), as an emerging QA task, enables innovative applications to effectively perceive, access, and understand complex biomedical knowledge [2] [3]. Automated question answering has made significant progress with multiple publicly available QA models, and datasets [4][5],[6]. However, in the clinical domain, the problem was relatively unexplored. Despite many efforts, biomedical QA still faces several key challenges such as dataset scaling, annotation, limited domain knowledge amongst data scientists, lack of explainability, limitations of performance evaluation methods, fairness, and bias [7]. For clinicians and their patients, diagnosis is central to management of a disease condition. All the patient data including history, physical exam, laboratory testing or consultations either supports generation of a diagnosis or is a result of further evaluation and management of the diagnosis. With the increasing adoption of electronic health record, the volume of patient data has been increasing at an exponential pace overwhelming the clinicians and leading to challenges with diagnosis and burnout. Thus, diagnosis specific QA models can help healthcare providers overcome these challenges by querying the electronic health record(EHR) efficiently and accurately, but the lack of diagnosis-specific QA datasets limits such model development. In this work, we have proposed a clinician validated pipeline for building QA datasets from EHR data which can be used to develop clinical QA models with better performance and explainability.

## 2 Methods

We have implemented our pipeline **(Figure1)** for building the QA datasets for the CVA which can be extensible for another diagnosis also. We used MIMIC-III data [8] which is a well-researched open source EHR database. We took ‘Noteevents’ data from the MIMIC-III. Notevents data has 2083180 clinical information files containing ‘Discharge summary’ ‘Echo’ ‘ECG’ ‘Nursing’ ‘Physician ‘ ‘Rehab Services’ ‘Case Management’ ‘Respiratory’ ‘Nutrition’ ‘General’ ‘Social Work’ ‘Pharmacy’ ‘Consult’ ‘Radiology’ ‘Nursing/other’. We used discharge summaries (n=59652) of MIMIC-III data for building a QA dataset. Discharge summary is an exhaustive source of patient information such as diagnosis, medications, procedures where medical information is summarised in multiple segments such as Chief Complaint, Major Surgical or Invasive Procedure, History of Present Illness, History of past illness, Discharge Diagnosis and Discharge medication. We used regular expressions to separate these sections.

**Figure1:**
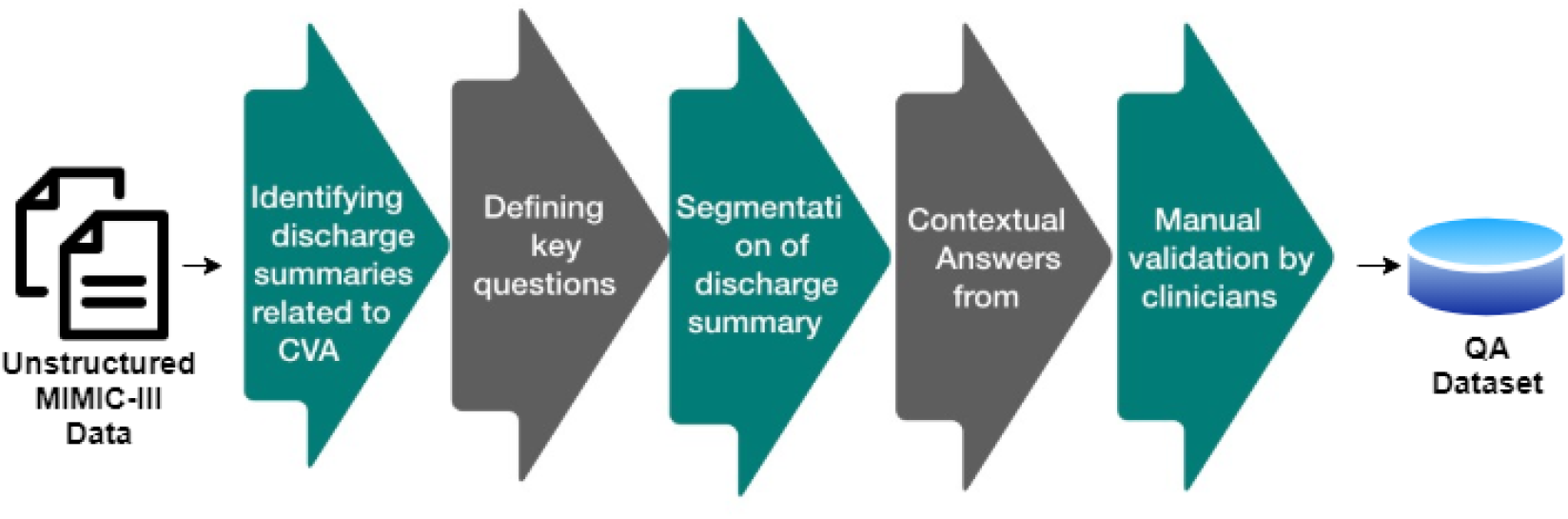
Flowchart of QA dataset developement pipeline.

### 2.1 Identification of Diagnosis from MIMIC-III discharge summaries

Identifying diagnoses of patients from the discharge summaries is challenging. Keywords search can be the easiest possible way to identify diagnosis. However, just keyword search from the entire discharge summary can generate false positives. In order to discard false positives, we reduced the keyword search space for the diagnosis to a single section ‘Discharge Diagnosis’. We further looked at the synonyms of CVA in the selected ‘Discharge Diagnosis’ section. Hence, we identified 568 CVA-related discharge summaries in the MIMIC-III discharge summary dataset.

### 2.2 Defining key questions for the dataset

Our team of five clinicians decided the six key questions **(Table 1)** which are commonly asked for the diagnosis and diverse aspects related to management of a patient with CVA. These questions were framed considering generalizability of their to other disease diagnoses.

**Table1:** Segmentation of discharge summary based on the question and related context.

| Questions | Context<br>Initial Segment | Context<br>End segment |
| --- | --- | --- |
| 1)Does the patient have CVA? | Discharge Diagnosis | Discharge Condition |
| 2)Was the patient discharged on Aspirin? | Discharge Medication | Discharge Diagnosis |
| 3)Is the patient on dialysis? | History of present illness | Past Medical History |
| 4)What is the patient's EF? | Echo results | Brief Medical Course |
| 5)Does the patient have heart failure? | Pertinent Results | Discharge Diagnosis |
| 6)Is there a CT available? | Discharge Diagnosis | Discharge Condition |

### 2.3 Finding contextual answers for a given question

Implementation of QA models can be a computationally expensive process. Therefore, we performed segmentation for the discharge summaries **(Table1)** in order to find the best context segment for a given question. We subsequently performed keyword searches to find the correct answer.

## 3 Results

Metric based evaluation can be challenging for QA models. Therefore, we had five clinicians perform manual validation for the 20%(112 CVA discharge summary) of the dataset. Validation was done in a binary way i.e. if QA pairs generated by pipeline were correct then we have reported the performance by 1 and if incorrect then we have reported performance by 0. Finally, the accuracy **(Figure2)** of the pipeline was calculated as a measure of correctness.

**Figure2:**
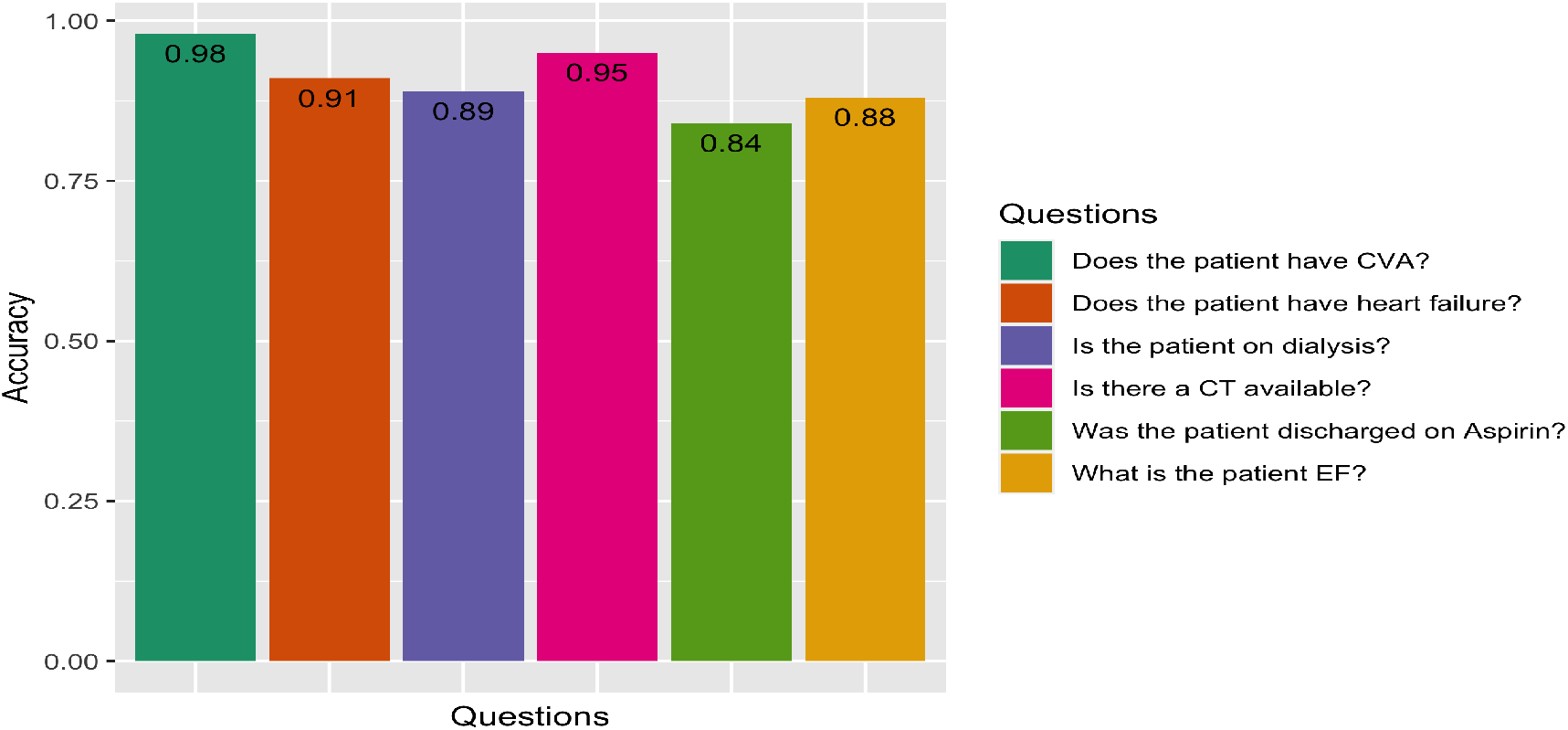
QA dataset pipeline performance based on clinician validation.

## 4 Discussion

In this paper, we curated a QA dataset for patients with encounter diagnosis of CVA from the MIMIC-III database discharge summaries, and have created a methodological pipeline which can possibly be generalized for other diagnoses too. Accuracy of our method in development of the QA dataset achieved an accuracy of 0.90(with 90% CI [0.82,0.99]) tested against the current gold standard of expert clinicians. While many of the questions that clinicians routinely interrogate the EHR for are common, keyword based search results which can miss clinical context are a challenge[9]. For,e.g., when performing a keyword search for a medication like Aspirin on discharge prescription, as in our use case, patients who have allergy to Aspirin will also return positive. Hence, methods such as segmentation of the context document as in our methodology helps with improving both computational efficiency and accuracy. Another common issue in development of QA data set for clinical needs is the variability of answers in the context[10]. In our dataset,while ejection fraction(EF) of the heart is mostly reported in numerical values, many discharge summaries mention the same as “ normal value”. Hence, for certain QA datasets multiple possible answers might need to be classified as a single answer. Generalizability and scalability of models in healthcare has been a significant challenge for AI due to limitations of available datasets[11]. For our future work, we plan on replicating our pipeline development methodology on a few other identified diagnoses using EHR datasets from different institutions. Our ultimate goal is to build a generalizable and scalable open-source tool for generation of diagnosis-specific ClinicalQA datasets and models which can be used by healthcare providers.

## 5 Conclusion

Our approach of developing semi-automated pipelines can help curate healthcare QA datasets is a unique approach developed in collaboration with clinicians. Utilizing our methodology we minimized the challenge of labor-intensive labeling, large-scale validation by clinicians, and improved computational efficiency with a high level of accuracy.

## Data Availability

Data is publicly available at https://physionet.org/content/mimiciii-demo/1.4/

